# Management of Engraftment Arrhythmias Associated with Human Induced Pluripotent Stem Cell-Derived Cardiomyocytes Transplantation

**DOI:** 10.64898/2026.01.09.26343817

**Authors:** Aixue Zhang, Rui Jing, Xiaocheng Liu, Yunqiang Zhang, Yuanlu Chen, Yongde Wang, Jianyong Wang, Zhipeng Guo, Jianliang Zhang, Qin Yang, Yufan Liu, Yanwei Wei, Yue Fu

**Affiliations:** TEDA International Cardiovascular Hospital, Tianjin University, Tianjin, China

**Keywords:** **Keyword**: Heart failure, Ischemic cardiomyopathy, iPS-Cardiomyocytes, Engraftment arrhythmias, Cardiac regeneration

## Abstract

**Background:** Human induced pluripotent stem cell-derived cardiomyocytes (hiPSC-CMs) represent a highly promising approach for cell-based replacement therapy in heart failure. However, the development of graft-related ventricular arrhythmias immediately following transplantation impedes its clinical translation. To date, there have been no reports worldwide characterizing the features of engraftment arrhythmias after hiPSC-CMs transplantation in human. Consequently, this study aims to analyze the characteristics of ventricular arrhythmias and the efficacy of antiarrhythmic drugs following hiPSC-CMs transplantation, and to identify risk factors associated with the occurrence of ventricular arrhythmias.

**Methods:** This study enrolled patients who underwent coronary artery bypass grafting (CABG) combined with hiPSC-CMs implantation under general anesthesia with cardiopulmonary bypass at our hospital between November 2023 and November 2025. Patients were assigned to low- and medium-dose groups based on the injected cardiomyocyte dose: 0.5×10⁸ cells and 1.5×10⁸ cells, respectively. Eleven patients were enrolled in each dose group. Ventricular tachycardia-related parameters were compared between the two groups after hiPSC-CMs implantation, and the characteristics of ventricular tachycardia episodes as well as the effectiveness of antiarrhythmic drugs were analyzed. A multivariate logistic regression model was applied to analyze risk factors influencing the occurrence of ventricular arrhythmias.

**Results:** No statistically significant differences were observed between the two groups in terms of gender, age, LVEF, LVEDD, myocardial infarction percentage, intraoperative CPB time, mean heart rate, QRS duration, QTc interval, PVC burden, or daily dose of beta-blocker or cell viability (P > 0.05). The incidence of VT was significantly higher in the medium-dose group compared to the low-dose group (P < 0.05). No statistically significant differences were observed between the groups regarding the time interval from hiPSC-CMs implantation to initial VT onset, the slowest frequency at initial VT onset, the fastest VT frequency, or VT duration (P > 0.05). Further analysis of VT in both groups at different time points after implantation revealed that the incidence of VT in the medium-dose group was significantly higher than that in the low-dose group on days 14, 21, and 28. However, comparisons of the fastest VT frequency at various time points and the incidence of VT at day 7 between the two groups showed no statistically significant differences (P > 0.05). Among all enrolled patients, VT occurred in 12 patients (54.5%). Based on ECG localization, the origin of VT in all cases was identified at the cell injection site. The time from hiPSC-CMs injection to initial VT onset ranged from 5 to 20 days (median 8.5 days). VT persisted for 9 to 411 days (median 59 days) before spontaneous termination, with 9 patients (75% of those with VT) experiencing VT lasting more than 1 month. The slowest frequency at initial VT onset ranged from 55 to 96 bpm (72.58 ± 9.86 bpm), while the fastest recorded frequency reached 111 to 185 bpm (146.33 ± 21.44 bpm). Hemodynamics remained stable in all patients during increases in VT frequency. Throughout the observed VT episodes, QRS morphology was consistently monomorphic, although cycle length varied. During VT episodes, overdrive pacing via the temporary epicardial atrial lead successfully suppressed but did not terminate VT. While overdrive pacing failed to reduce the frequency of VT, it provided a critical window for optimizing antiarrhythmic drug therapy to control VT. Cardioversion also failed to terminate VT. Administration of beta-blockers, ivabradine, or amiodarone controlled the VT frequency to a range of 55-95 bpm (79.67 ± 12.51 bpm) but likewise did not terminate the arrhythmia. VT terminated spontaneously in patients, after which it either did not recur, recurred intermittently, or reoccurred as sustained VT after a period of time. Multivariate logistic regression analysis indicated that the dose of hiPSC-CMs injection was an independent influencing factor for the risk of VT onset (P < 0.05).

**Conclusion:** The ectopic arrhythmia (EA) is primarily driven by an automaticity mechanism. It is characterized by early onset (median 8.5 days) and prolonged duration (median 59 days), with the cell injection dose identified as an independent risk factor (OR=9.00). Within the controlled dose range (0.5-1.5×10⁸ cells) and under strict clinical management, this type of arrhythmia can be effectively monitored and managed.

---

Heart failure affects over 1.2 billion people annually and represents the leading global cause of mortality and hospitalization ^[1]^. During myocardial infarction, approximately one billion cardiomyocytes are permanently lost. Among survivors, persistent post-infarction myocardial deficits lead to chronic mechanical overload, which in turn triggers pathological remodeling of the surviving myocardium and further impairs contractile function. Ultimately, nearly 20% of patients subsequently progress to heart failure ^[2,3]^. Despite a severe deficiency of donor organs, heart transplantation remains the gold-standard therapy for end-stage heart failure patients ^[4]^. Human induced pluripotent stem cell-derived cardiomyocytes (hiPSC-CMs) offer a highly promising strategy for cell replacement therapy following myocardial infarction. However, the occurrence of engraftment arrhythmias post-transplantation poses a significant barrier to clinical translation. Previous clinical studies have reported transient but severe arrhythmias following hiPSC-CMs transplantation in large animal models, such as non-human primates and pigs ^[5–8]^. To date, there have been no global reports characterizing transplantation-associated ventricular arrhythmias following hiPSC-CMs transplantation in humans. Therefore, this study aims to analyze the characteristics of ventricular arrhythmias and the efficacy of antiarrhythmic drugs after hiPSC-CMs transplantation, and to identify risk factors influencing their occurrence.

## Materials and Methods

### General Information

This study enrolled a total of 22 patients who underwent coronary artery bypass grafting (CABG) combined with hiPSC-CMs implantation under general anesthesia with cardiopulmonary bypass at our hospital between November 2023 and November 2025. The cohort consisted of 20 males (90.9%), with ages ranging from 41 to 73 years (mean ± SD: 59.05 ± 8.38 years).

Inclusion Criteria: Age ≥ 18 years at the time of providing written informed consent; Evidence of myocardial infarction in the left anterior descending (LAD) coronary artery territory, as confirmed by nuclear myocardial perfusion/metabolism imaging or cardiac magnetic resonance (CMR); Left ventricular ejection fraction (LVEF) ≤ 40% as measured by CMR; Planned for CABG surgery. Exclusion Criteria: History of pacemaker, implantable cardioverter-defibrillator (ICD), or cardiac resynchronization therapy (CRT) device implantation; Diagnosis of malignancy within the past 5 years; Presence of autoimmune diseases; History of organ transplantation; Scheduled for concurrent other cardiac surgical procedures (excluding ventricular aneurysmectomy or left atrial appendage exclusion/occlusion); History of malignant ventricular arrhythmias; contraindications to CABG surgery; Inability to undergo required CMR or PET/CT examinations; Contraindications to immunosuppressive therapy; Female patients who were pregnant, breastfeeding, or had a positive pregnancy test.

The study protocol was approved by the Institutional Ethics Committee of TEDA International Cardiovascular Hospital (Approval No.: 2023IIY05; Project ID: 202308). All participants provided written informed consent prior to enrollment.

## Methods

Patients’ demographic data were systematically collected after consensus. Preoperative Electrocardiogram (ECG) parameters: QRS duration and corrected QT (QTc) interval. Preoperative ambulatory ECG parameters: mean heart rate and PVC burden. Preoperative CMR data: LVEF, LVEDD, and percentage of myocardial infarction. Preoperative metoprolol tartrate usage, intraoperative CPB time, and other relevant parameters.

### Surgical Procedure

Ultra-purified hiPSC-CMs were manufactured by HELP Therapeutics. Eligible patients were assigned into low- and medium-dose groups, with an injection dose at 0.5 × 10⁸ cells and 1.5 × 10⁸ cells, respectively. Eleven patients were enrolled in each group.

Transplantation of hiPSC-CMs was performed during a cardiopulmonary bypass surgery under general anesthesia. The target injection site was identified as the border zone between infarcted and viable myocardium. This region was mapped preoperatively using CMR with tissue characterization (Mapping) in conjunction with Nuclear myocardial perfusion/metabolism imaging. A multi-point injection technique (Figure 1) was employed. Injections were administered sequentially from the endocardial side (within the inner one-third of the ventricular wall thickness) toward the epicardial side. Postoperatively, a one-year immunosuppressive regimen was initiated, primarily consisting of enteric-coated mycophenolate sodium and tacrolimus. Tacrolimus trough levels were maintained within a target range of 5-10 ng/ml.

**Figure 1.**
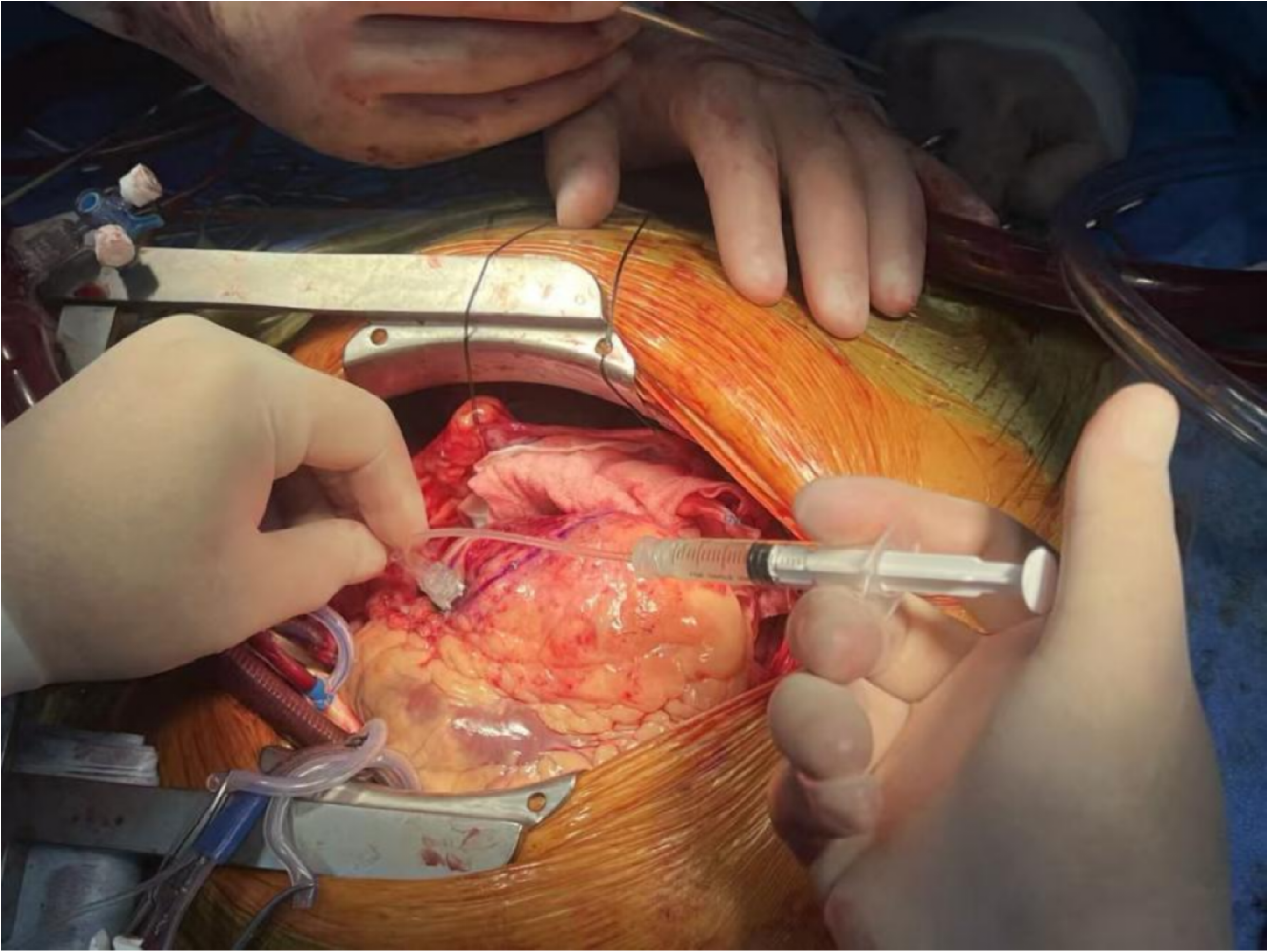
A multi-point injection technique of Intraoperation

### Antiarrhythmic Medications

Prior to implantation, beta-blockers were routinely administered to both patient groups, with no statistically significant difference in daily dosage (Table 1). One patient in each group received oral amiodarone for the treatment of premature Ventricular Complex (PVC). In the low-dose group, amiodarone was prescribed at 0.2 g twice daily. In the medium-dose group, amiodarone was initiated at 0.2 g three times daily, followed by weekly dose adjustments, each involving a reduction of the total daily dose by 0.2 g. Through this stepwise tapering process, the maintenance dose was ultimately reduced to 0.2 g once daily. Aside from these two medications, no other antiarrhythmic drugs were used in either group.

**Table 1.** Comparison of Baseline Characteristics Between the Low-dose and Medium-dose GroupsDose-Dependent VT.

|  | Low-dose<br>(n=11) | Medium-dose<br>(n=11) | P value |
| --- | --- | --- | --- |
| Male {n (%)} | 10 (90.9) | 10 (90.9) | 1.000 |
| Age (years) | 57.00±9.11 | 60.92±6.80 | 0.240 |
| LVEF (%) | 31.73±6.44 | 28.08±2.81 | 0.077 |
| LVEDD (mm) | 60.45±6.01 | 60.85±7.24 | 0.888 |
| Myocardial infarction<br>percentage (%) | 22.02±7.57 | 21.25±8.28 | 0.822 |
| Intraoperative CPB CABG<br>time (h) | 127.18±40.66 | 124.46±30.84 | 0.854 |
| Mean heart rate (bpm) | 72.27±8.96 | 71.36±9.15 | 0.748 |
| QRS duration (ms) | 99.91±12.56 | 101.73±16.87 | 0.777 |
| QTc interval (ms) | 439.36±16.03 | 434.91±21.67 | 0.590 |
| PVC burden (%) | 1.57±4.82 | 0.65±1.69 | 0.556 |
| Daily dose of beta-blocker<br>(mg) | 29.55±10.11 | 27.27±12.27 | 0.562 |
| Cell viability (%) | 79.46% ± 1.95% | 80.19% ± 1.58% | 0.347 |

### Atrial Pacing

Following the implantation of hiPSC-CMs, a conventional temporary atrial epicardial pacing lead was placed in all patients. If the patient developed a high-rate (>100 bpm) or sustained Engraftment arrhythmia (EA), atrial overdrive pacing was administered, with the pacing rate set 5–10 bpm above the EA frequency. Effective pacing was defined as a reduction in EA burden to below 20% within 24 hours.

### Electrophysiology Assessment

EA was monitored via 12-lead ECG and 12-lead ambulatory electrocardiogram (Holter), acquired respectively from a multi-lead ECG machine and an ambulatory ECG monitor. The multi-lead ECG machine was manufactured by Shenzhen Edan Instruments, Inc., with the model number SE-1201, while the ambulatory ECG monitor was produced by Shenzhen Medical Instrument Technology Co., Ltd., with the model number BI6812. Routine 12-lead ECG examinations were performed on days 1–7, 14, 21, and 28 post-implantation. 12-lead Holter monitoring was conducted on days 1, 3, 5, 7–14, 16, 19, 22, 25, and 28. The frequency of 12-lead Holter could be increased as clinically warranted in cases of EA.

EA, including PVC and VT, were compared between the two groups following hiPSC-CMs implantation. These included the incidence of VT, the slowest frequency at initial VT onset, the fastest recorded VT frequency, and the duration of VT episodes.

The characteristics of VT episodes and the efficacy of antiarrhythmic drugs were analyzed. VT characteristics encompassed: the time interval from hiPSC-CMs implantation to initial VT onset, the morphology (monomorphic/polymorphic), the slowest frequency at onset, the fastest recorded frequency, and the episode duration. The evaluated antiarrhythmic drugs included beta-blockers, amiodarone, and ivabradine.

### Statistical Analysis

Data were processed using SPSS software (version 29.0). The Shapiro-Wilk test was applied to assess the normality of continuous variables, and the Levene’s test was used to evaluate the homogeneity of variances. Continuous data are presented as mean ± standard deviation or median [M], as appropriate. Categorical data are expressed as number of cases (percentage). For between-group comparisons, continuous variables were analyzed using the independent samples t-test or the Mann-Whitney U test, based on their distribution. Categorical variables were compared using the chi-square (x²) test. A logistic regression model was employed for multivariable analysis, incorporating all covariates considered clinically relevant. The model yielded odds ratios (OR) with corresponding 95% confidence intervals (CI). A two-sided P-value of less than 0.05 was considered statistically significant.

## Results

### Baseline Characteristics

No statistically significant differences were observed between the two groups in terms of gender, age, LVEF, LVEDD, myocardial infarction percentage, intraoperative CPB time, mean heart rate, QRS duration, QTc interval, PVC burden, or daily dose of beta-blocker or cell viability (P > 0.05), as detailed in Table 1.

The incidence of VT was significantly higher in the medium-dose group compared to the low-dose group (P < 0.05). No statistically significant differences were observed between the groups regarding the time interval from hiPSC-CMs implantation to initial VT onset, the slowest frequency at initial VT onset, the fastest VT frequency, or VT duration (P > 0.05), as detailed in Table 2.a. Further analysis of VT in both groups at different time points after implantation revealed that the incidence of VT in the medium-dose group was significantly higher than that in the low-dose group on days 14, 21, and 28. However, comparisons of the fastest VT frequency at various time points and the incidence of VT at day 7 between the two groups showed no statistically significant differences (P > 0.05), as detailed in Table 2.b and Line Chart 1.

**Table 2.**
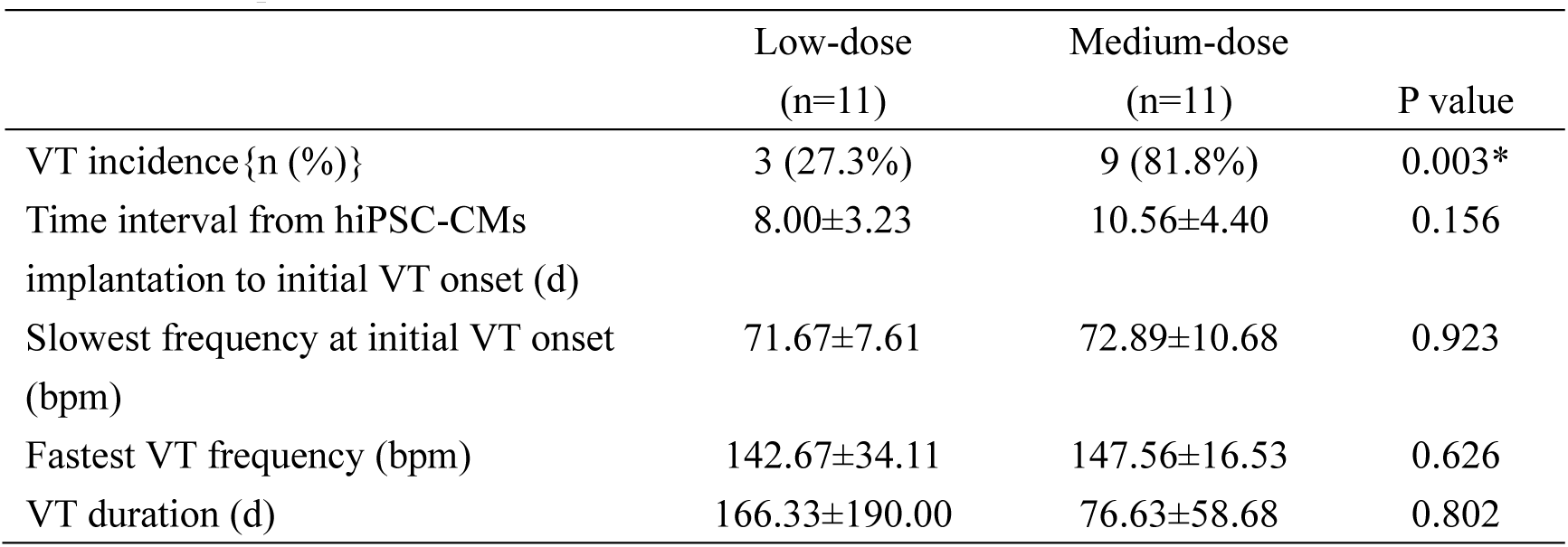
a Comparison of VT Characteristics Between the Low- and Medium-dose.

|  | Low-dose<br>(n=11) | Medium-dose<br>(n=11) | P value |
| --- | --- | --- | --- |
| VT incidence{n (%)} | 3 (27.3%) | 9 (81.8%) | 0.003* |
| Time interval from hiPSC-CMs<br>implantation to initial VT onset (d) | 8.00±3.23 | 10.56±4.40 | 0.156 |
| Slowest frequency at initial VT onset<br>(bpm) | 71.67±7.61 | 72.89±10.68 | 0.923 |
| Fastest VT frequency (bpm) | 142.67±34.11 | 147.56±16.53 | 0.626 |
| VT duration (d) | 166.33±190.00 | 76.63±58.68 | 0.802 |

**Table 2.**
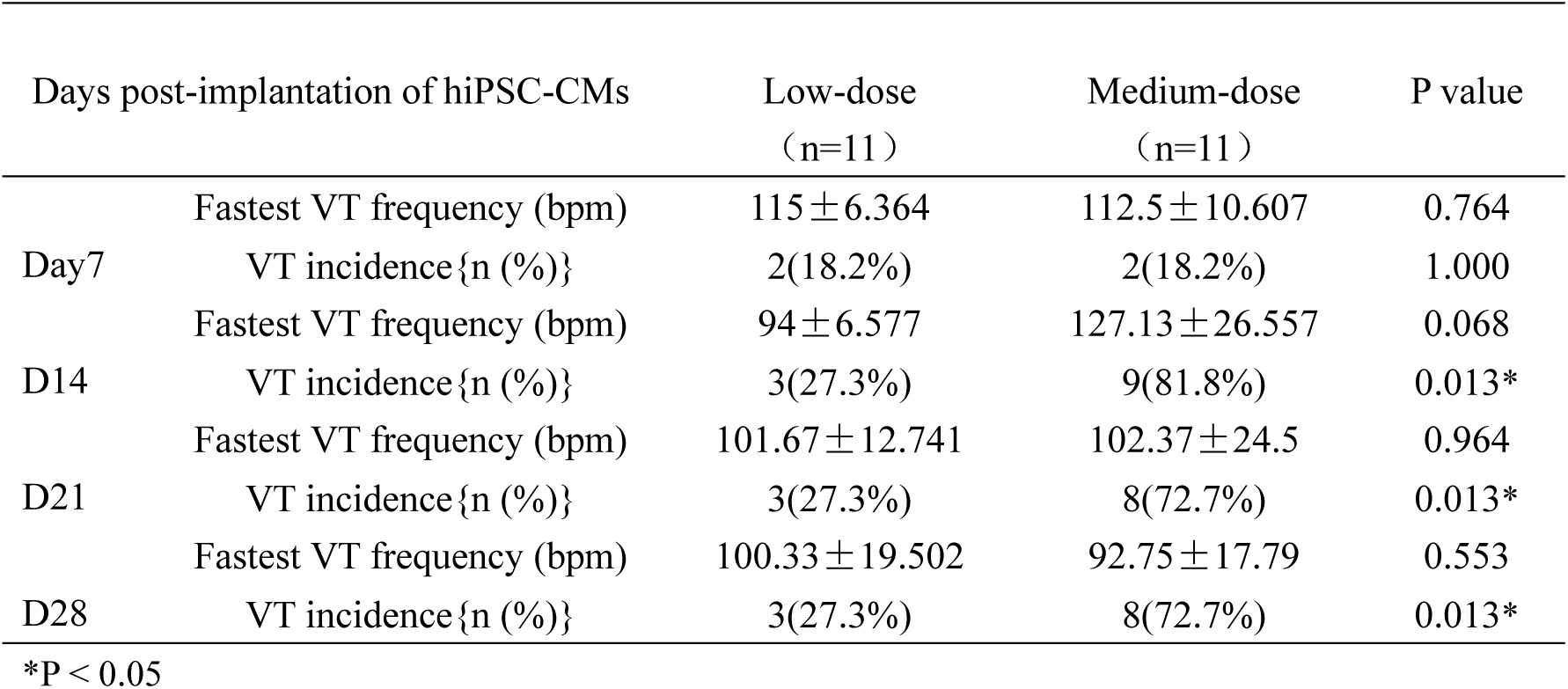
b. Comparison of VT Characteristics Between Low- and Medium-dose.

Line Chart 1. Incidence of VT at different time points post-implantation

### Characteristics of VT Episodes

Among all enrolled patients, VT occurred in 12 patients (54.5%). ECG localization identified the origin of VT in all cases at the cell injection site (Figures 2a,b). The time from hiPSC-CMs injection to initial VT onset ranged from 5 to 20 days (median 8.5 days). The VT initially presented as paroxysmal episodes. Over a period of several hours, the frequency of VT episodes progressively increased until evolving into a persistent VT state (Figure 3 and Figure 4 ). The slowest frequency at initial VT onset ranged from 55 to 96 bpm (mean ± SD: 72.58 ± 9.86 bpm), while the fastest recorded VT frequency reached 111 to 185 bpm (mean ± SD: 146.33 ± 21.44 bpm). Hemodynamic stability was maintained in all patients during periods of increasing VT frequency. Throughout all observed VT episodes, QRS morphology remained monomorphic, although cycle length variability was noted. During VT episodes, overdrive pacing via the temporary epicardial atrial lead successfully suppressed but did not terminate VT. While overdrive pacing failed to reduce the frequency of VT, it provided a critical window for optimizing antiarrhythmic drug therapy to control VT in the patient.(Figure 5). Cardioversion also failed to terminate the arrhythmia. Administration of beta-blockers, ivabradine, or amiodarone controlled the VT frequency to a range of 55-95 bpm (mean ± SD: 79.67 ± 12.51 bpm) but likewise did not achieve termination. Details of patient-specific antiarrhythmic drug use are presented in Table 3. VT eventually terminated spontaneously in patients. Following termination, VT either did not recur, recurred intermittently, or reoccurred as sustained VT after a variable period. After discharge, patients were provided with a Cardio-Guardian event monitor for continued tracking of VT persistence. Analysis of the monitor data revealed that VT persisted for 9 to 411 days (median 59 days) before spontaneous termination. Among patients with VT, 9 patients (75%) experienced VT lasting more than 1 month.

**Figures 2.**
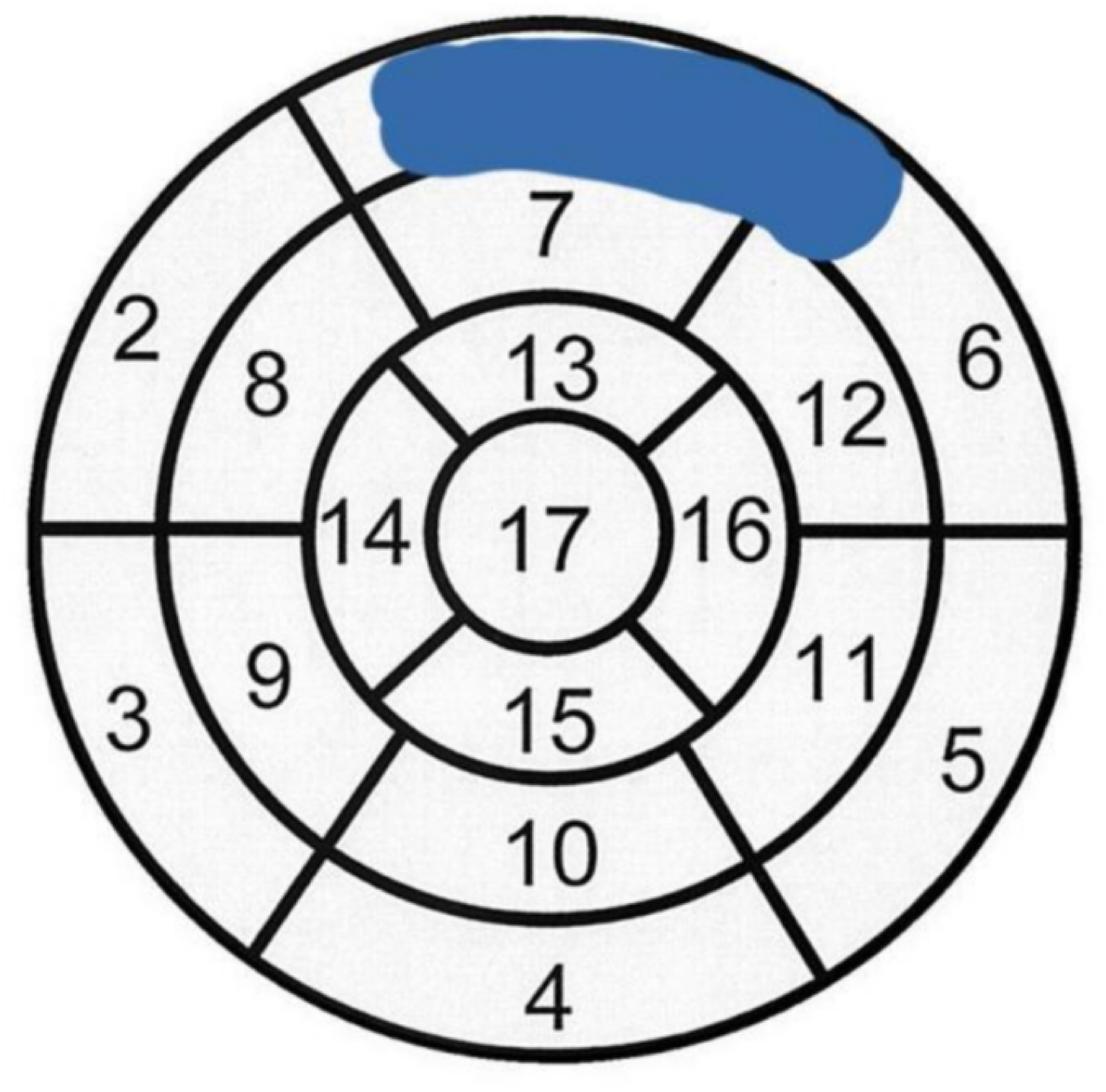
(a,b). Injection sites of hiPSC-CMs and corresponding locations of surface ECG leads.

**Figures 2.**
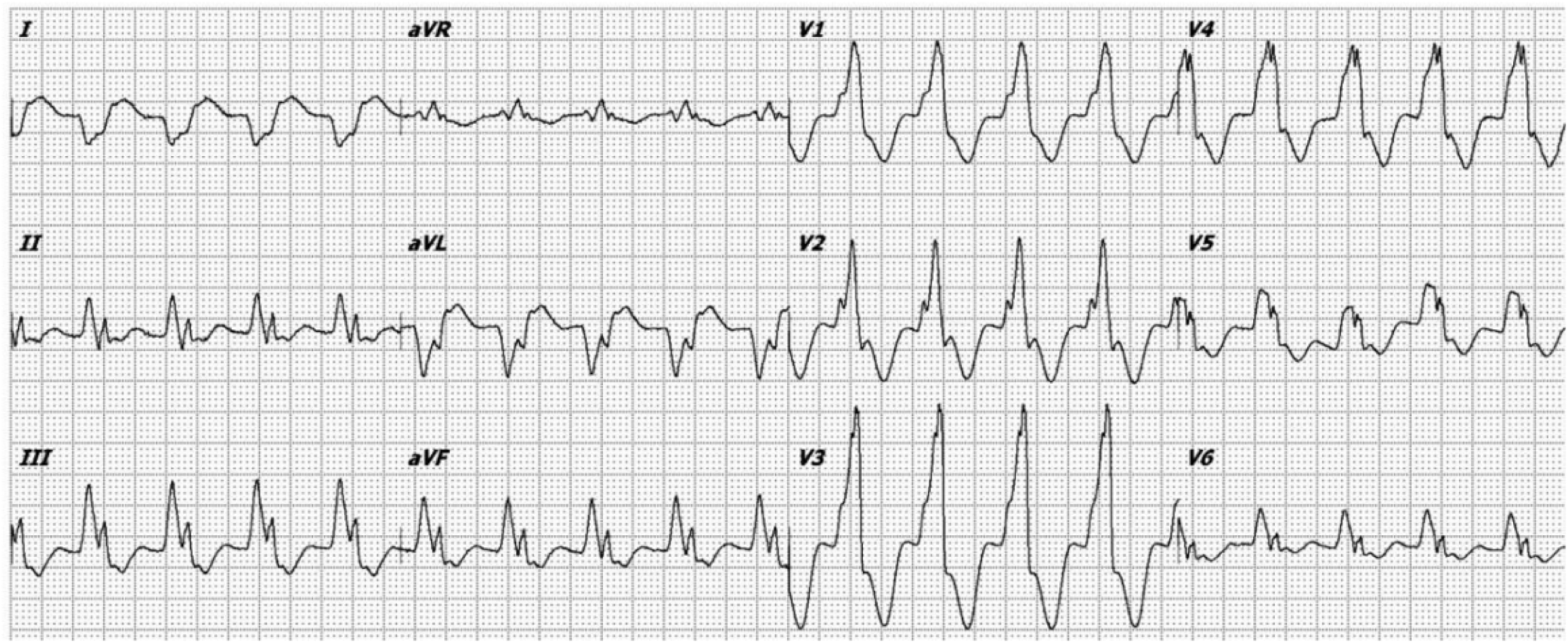
a. Injection sites of hiPSC-CMs.

**Figures 2.**
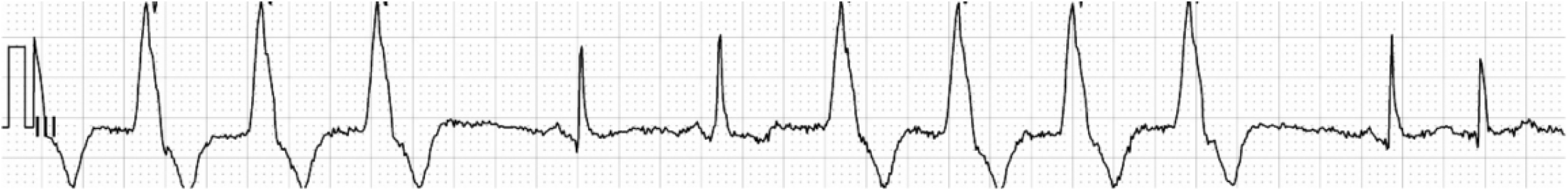
b. Manifestation of surface ECG leads related to the cell injection site.

**Figure 3.**
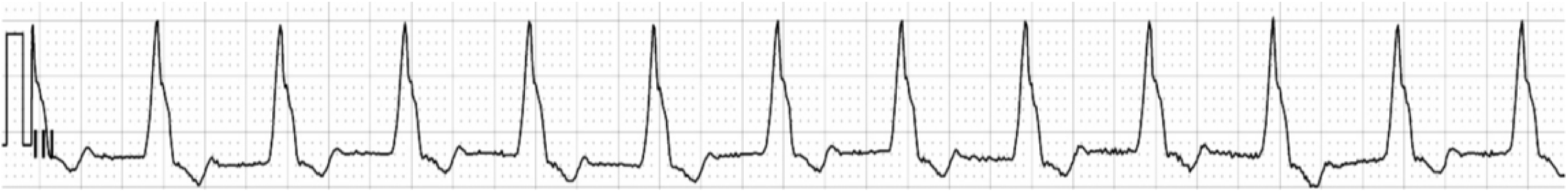
(a,b). Evolution of VT in low-dose patient group.

**Figure 3.**
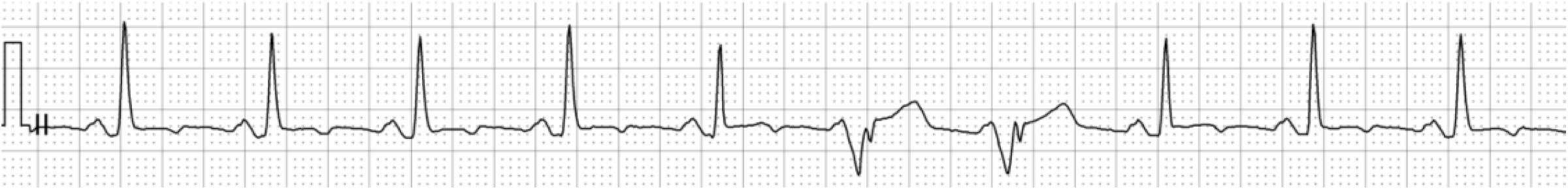
a. ECG manifestations during initial VT occurrence.

**Figure 3.**
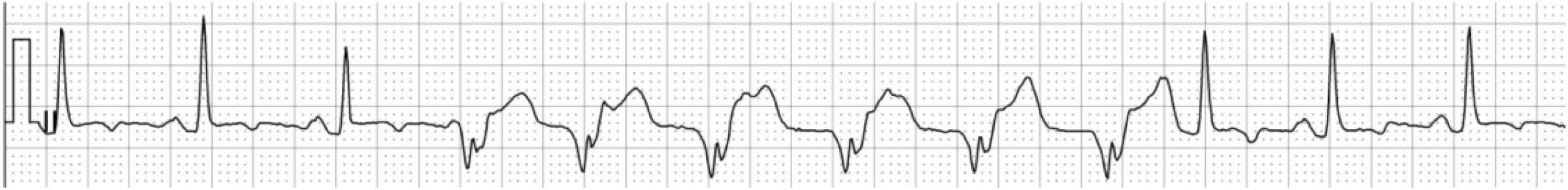
b. 2 hours later.

**Figure 4.**
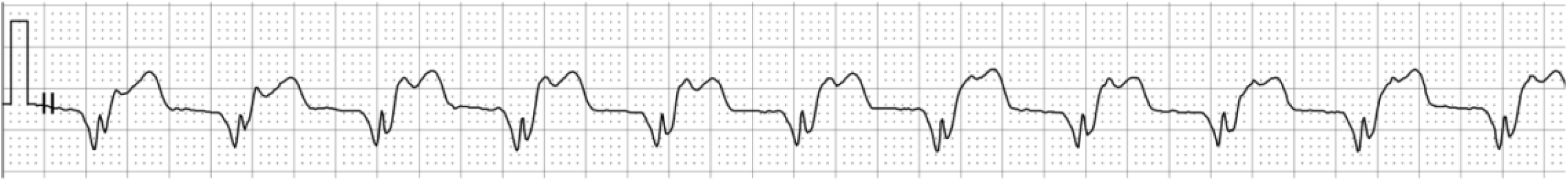
(a,b,c). Evolution of VT in Medium-dose patient group.

**Figure 4.**
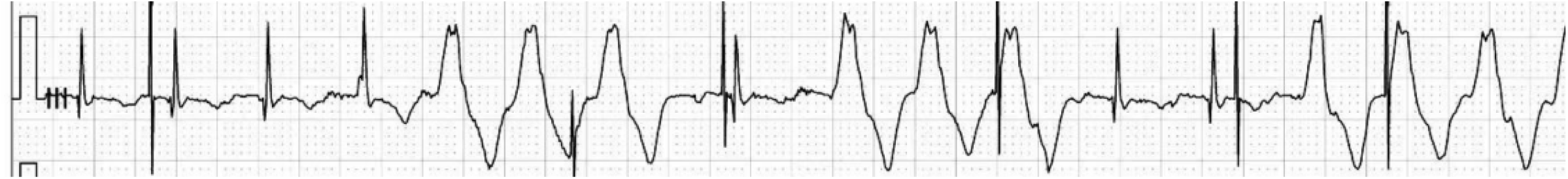
a. ECG manifestations during initial VT occurrence.

**Figure 4.**
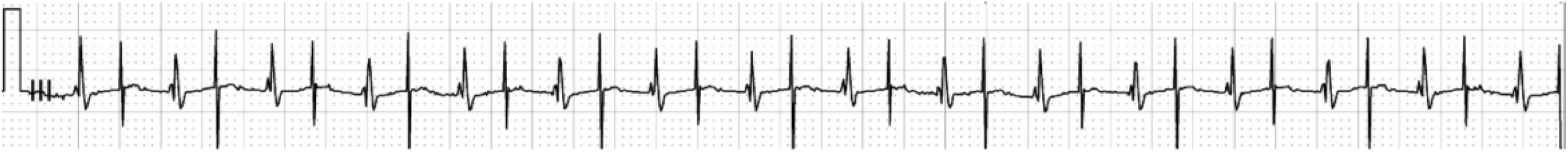
b. 20 hours later.

**Figure 4.**
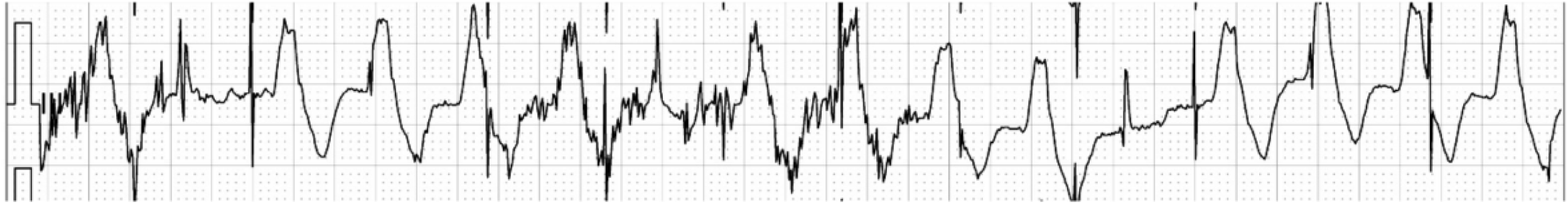
c. 22 hours later.

**Figures 5.**
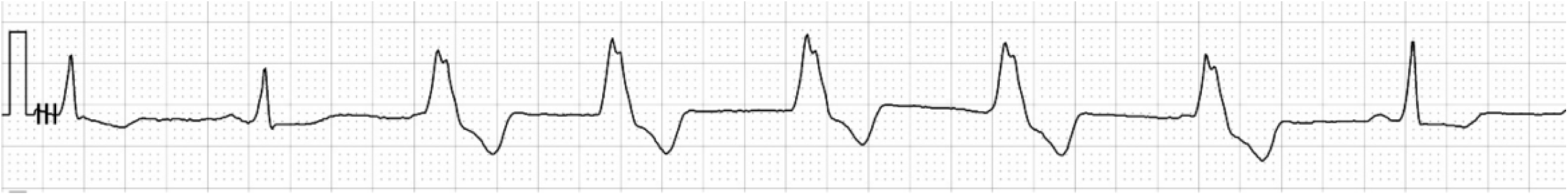
(a,b,c,d). ECG Manifestations of Atrial Overdrive Pacing Suppression.

**Figures 5.a.**
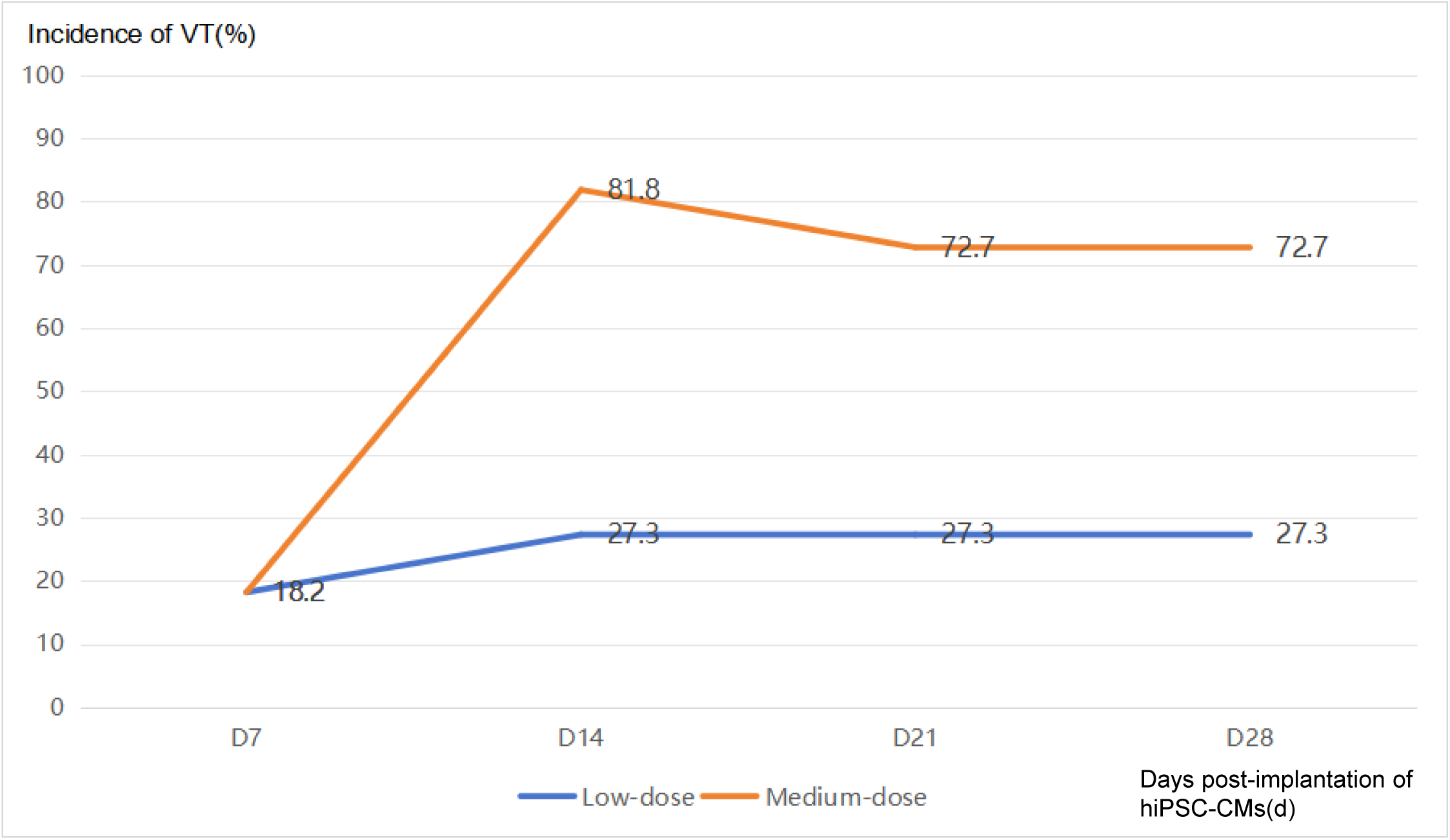
Overdrive Pacing Therapy b. Overdrive Pacing:Effective c. 4 hours later Overdrive pacing failed to reduce the frequency of VT; The dose of β-blocker was increased and ivabradine was added. d. 8 hours later The frequency of VT was effectively controlled.

### Multivariate Logistic Regression Analysis of Risk Factors for VT

A multivariate logistic regression model was employed to analyze the association between the occurrence of VT and 9 predictors: preoperative QRS duration, QTc interval, preoperative PVC burden, daily dose of metoprolol tartrate, preoperative LVEF and LVEDD, preoperative myocardial infarction percentage, intraoperative CPB time, and dosage of hiPSC-CMs. All statistically significant variables from the univariate analyses were subsequently entered into a multivariate logistic regression model. The results identified the dose of hiPSC-CMs injection as an independent influencing factor for the risk of VT onset (P < 0.05). Detailed results are presented in Table 5.

**Table 4.** Antiarrhythmic Drug Therapy in Patients with VT.

| Drug Regimen | Number of VT Patients Receiving Therapy {n (%)} |
| --- | --- |
| Beta-blocker + Ivabradine + Amiodarone | 3 (25) |
| Beta-blocker + Ivabradine | 5 (41.7) |
| Beta-blocker + Amiodarone | 1 (8.3) |
| Beta-blocker | 3 (25) |

**Table 5.**
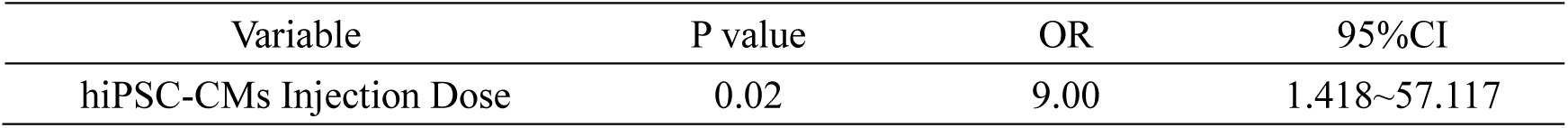
Results of Multivariate Logistic Regression Analysis on the Risk of VT Episodes.

| Variable | P value | OR | 95%CI |
| --- | --- | --- | --- |
| hiPSC-CMs Injection Dose | 0.02 | 9.00 | 1.418~57.117 |

## 3 Discussion

The advent of hiPSC-CMs in 2007 opened new avenues for myocardial regeneration therapy, establishing them as a highly promising regenerative strategy for addressing irreversible myocardial damage following infarction. This therapeutic approach aims to replace cardiomyocytes lost during myocardial infarction through the implantation of exogenous cells ^[9,10]^. Numerous studies utilizing clinically relevant large animal models have demonstrated that while this therapy holds potential for improving cardiac function, it is also associated with ventricular arrhythmias – termed EA – which represent a foreseeable and common complication following intramyocardial injection of hiPSC-CMs ^[5–7,11,12]^.

Multiple animal studies have indicated that EA typically emerges within the first week post-cardiomyocyte transplantation ^[5–7,12,13]^. The initial presentation often involves repetitive runs of non-sustained VT, which generally progresses to sustained VT, frequently exhibiting a polymorphic morphology ^[7]^. In the present study, EA occurred between 5 and 20 days post-transplantation, with a median of 8.5 days. The arrhythmia initially manifested as paroxysmal episodes. Over a period of several hours, the frequency of VT episodes progressively increased until evolving into a persistent VT state, which is largely consistent with the findings described above. Observation throughout the EA episodes in our cohort revealed a monomorphic VT pattern. This is likely attributable to the predominance of a single, high-frequency automatic focus originating from the engrafted cardiomyocyte cluster.

In contrast to ventricular arrhythmias caused by scar-mediated reentrant circuits—which typically exhibit fixed cycle lengths and can be terminated by rapid pacing or external cardioversion—Dinesh Selvakumar et al. ^[13]^ analyzed EA in porcine models following hiPSC-CMs implantation and identified enhanced automaticity as the underlying mechanism. Their conclusions were supported by several key observations: 1) Subsequent electroanatomic mapping performed four weeks post-transplantation localized the origin of EA to the focal injection site, a finding later confirmed by histological analysis. 2) VT occurred spontaneously or was induced by catecholamine administration, but could not be initiated by programmed electrical stimulation; it terminated spontaneously but often restarted shortly thereafter. 3) Neither rapid ventricular pacing nor external cardioversion could restore sinus rhythm. 4) Cycle length variability was consistently observed. 5) During entrainment pacing attempts, the arrhythmia accelerated without demonstrating progressive fusion or resetting of the tachycardia cycle. In the present study, ECG localization confirmed that all VT originated from the cell injection sites. Overdrive pacing via the atrial lead successfully suppressed but did not terminate VT, and cardioversion was largely ineffective. VT terminated spontaneously, after which it either did not recur, recurred intermittently, or reoccurred as sustained VT after a period of time, with documented cycle length variability. Therefore, the characteristics of EA observed in our study are consistent with an automaticity-based mechanism. The automaticity of hiPSC-CMs possesses a dual nature. On one hand, it serves as a definitive macroscopic marker confirming successful phenotypic differentiation into cardiomyocytes ^[14]^. On the other hand, it is also a manifestation of cardiomyocyte immaturity ^[15]^.

Previous animal studies have shown that the resolution of EA coincides with the in vivo maturation process of hiPSC-CMs ^[5,11]^, typically occurring around one month after onset ^[5–7,12,13]^. This temporal correlation suggests that the immaturity of hiPSC-CMs may be a key determinant of their arrhythmogenicity ^[16]^. However, not all transplanted hiPSC-CMs achieve complete "electrical maturation", it is recognized that current hiPSC-CMs differentiation protocols yield a heterogeneous cell population comprising a mixture of ventricular-like, atrial-like, and pacemaker-like cardiomyocytes ^[17]^. In the present study, the duration of EA ranged from 9 to 411 days, with a median of 59 days. Notably, EA persisted for more than 1 month in 9 patients, accounting for 75% of all EA cases. This duration is substantially longer than that reported in preclinical animal models. This discrepancy is likely attributable to a combination of factors: the pathological nature of the host heart, the quantity and inherent properties of the transplanted cells, and the complex, dynamic integration process between them. Unlike previous studies based on healthy or acute injury animal models, the ischemic cardiomyopathy hearts in our patient cohort represented a chronically remodeled pathological microenvironment characterized by both electrical and structural remodeling. Extensive fibrosis, abnormal gap junction distribution, and chronic neurohormonal activation collectively create a substrate that is unfavorable for rapid and uniform electrophysiological integration. Concurrently, despite achieving high purity prior to transplantation, once implanted into this pathological milieu, the grafted cells are subjected to heterogeneous local stresses, variable blood supply, and non-uniform host cell contacts. These conditions can further exacerbate phenotypic divergence and functional heterogeneity within the cell population, potentially giving rise to or amplifying cellular heterogeneity. The protracted duration of EA observed in our study fundamentally underscores the clinical complexity and prolonged nature of the transition from mere "cell injection" to true "functional integration". Consequently, future therapeutic strategies must extend beyond optimizing pre-transplantation cell purity to actively guiding the post-engraftment fate and maturation of these cells in vivo. The overarching objective should be to facilitate the development of an electrically homogeneous, stable, and seamlessly integrated tissue graft with the host myocardium.

Emerging evidence confirms the presence of a fully functional β-adrenergic system within hiPSC-CMs ^[18]^. Furthermore, hiPSC-CMs exhibit distinct ion channel expression profiles compared to adult human ventricular cardiomyocytes (hV-CMs). Notably, hiPSC-CMs express two ionic currents that are absent in healthy adult hV-CMs: the hyperpolarization-activated cyclic nucleotide–gated channel current (I_f_) ^[19]^ and the T-type calcium channel current (I_CaT_) ^[20]^. Additionally, the expression level of the inward rectifier potassium current (I_K1_) in hiPSC-CMs is comparable to, or even lower than, that in adult human atrial cardiomyocytes (hA-CMs) ^[21]^, with some studies reporting its complete absence ^[22]^. In a voltage-clock model, the balance between I_K1_ and I_f_ is a key determinant of the inherent automaticity observed in hiPSC-CMs. Based on these electrophysiological characteristics, researchers including Dinesh Selvakumar ^[13]^ and Kenta Nakamura ^[7]^ have employed a combination of ivabradine and amiodarone to target EA. This combined regimen has been shown to effectively inhibit automaticity, reduce heart rate, and decrease EA burden. In this study, a total of four antiarrhythmic regimens involving three drugs—beta-blockers, ivabradine, and amiodarone—used either alone or in combination were evaluated. Given the patients’ underlying ischemic cardiomyopathy and the known electrophysiological properties of hiPSC-CMs, a low-to-moderate dose beta-blocker was maintained routinely both before and after transplantation. In three patients (25%), beta-blocker monotherapy provided adequate control of EA frequency. The most commonly employed antiarrhythmic strategy was the combination of a beta-blocker with ivabradine, which was used in 41.7% of EA patients. Triple therapy (beta-blocker + ivabradine + amiodarone) was administered in 25% of cases, while the combination of a beta-blocker with amiodarone was used in only one patient (8.3%). Assessment based on EA frequency control demonstrated that all four regimens achieved the goal of rate control. However, none were able to terminate EA, a finding consistent with previous research ^[13]^.

The controllable clinical phenotype of ventricular arrhythmias observed in our study stands in sharp contrast to the lethal outcomes reported in some large animal models. For instance, Kenta Nakamura et al. ^[7]^ transplanted up to 5×10⁸ embryonic stem cell-derived cardiomyocytes in porcine myocardial infarction models, observing significant arrhythmogenicity and a consequent high mortality rate. In contrast, the cell doses employed in our clinical trial ranged from 0.5 to 1.5 ×10⁸ cells. Notably, all VT episodes remained hemodynamically stable, were effectively controlled with pharmacological intervention, and did not progress to malignant transformation or result in direct mortality. A critical finding of our study is the statistical confirmation that cell dose is a pivotal determinant of arrhythmic risk. Multivariate logistic regression analysis identified the hiPSC-CMs injection dose as an independent risk factor for ventricular arrhythmia occurrence (P = 0.02), with an OR as high as 9.00 (95% CI: 1.418–57.117). This indicates that, compared to the low-dose group, the medium-dose group carried an approximately ninefold increased risk of developing ventricular arrhythmias.

In summary, this study provides the first systematic clinical characterization of ventricular arrhythmias following hiPSC-CMs transplantation. The results demonstrate that the EA is primarily driven by an automaticity mechanism. It is characterized by early onset (median 8.5 days) and prolonged duration (median 59 days), with the cell injection dose identified as an independent risk factor (OR=9.00). Within the controlled dose range (0.5-1.5×10⁸ cells) and under strict clinical management, this type of arrhythmia can be effectively monitored and managed.

This study has several limitations. First, the relatively small overall sample size may limit statistical power and hinder a comprehensive assessment of rare adverse events. Second, as a single-center observational study, potential biases in patient selection and treatment protocols cannot be excluded. Third, the inference regarding the arrhythmia mechanism is primarily based on clinical and electrophysiological observations, lacking direct electrophysiological evidence at the cellular or tissue level. Future research should involve multi-center, large-sample randomized controlled trials. These studies should incorporate advanced imaging and detailed electrophysiological mapping techniques to more comprehensively evaluate the risk-benefit profile of this therapeutic approach.

## Author Contributions

A Z was responsible for data synthesis, analysis, and manuscript drafting. J R was responsible for study design, manuscript revision, and finalization of the manuscript. X C contributed by providing conceptual guidance on the study design and performing intraoperative hiPSC-CMs injections. Y Z, Y C, Y W, and J W were responsible for analyzing electrocardiographic data and guiding the application of antiarrhythmic medications. Z G, and J Z were responsible for localizing hiPSC-CMs injection sites and performing CABG surgery. Q Y assisted in literature research and data collection. Y L and Y W collected clinical data. Y F were responsible for the acquisition of ECG data. All authors have participated sufficiently in the work. All authors contributed to editorial changes in the manuscript. All authors read and approved the final manuscript. All authors agree to be accountable for all aspects of the work in ensuring that questions related to the accuracy or integrity of any part of the work are appropriately investigated and resolved.

## Funding

This work was supported by the Funded by Tianjin Key Medical Discipline Construction Project under Grant No. TJYXZDXK-3-035C.

## Conflict of Interest

The authors declare no conflict of interest.

## Data Availability

The data that support the findings of this study are not publicly available due to patient privacy protection, but are available from the corresponding author upon reasonable request. The use of these data has been approved by the Ethics Committee of TEDA International Cardiovascular Hospital (approval number: [2023IIY05]).

## Notes

### Competing Interest Statement

The authors have declared no competing interest.

### Clinical Trial

the URL:http://www.chinadrugtrials.org.cn/clinicaltrials.searchlistdetail.dhtml; ID: CTR20232197

### Author Declarations

The study protocol was approved by the Institutional Ethics Committee of TEDA International Cardiovascular Hospital (Approval No.: 2023IIY05).

